# A Qualitative Study Investigating Sex Differences in the Needs and Priorities of Lower Limb Prosthesis Users

**DOI:** 10.64898/2026.11.20.26368208

**Authors:** Tess M. R. Carswell, Helen Monkman, Joshua W. Giles

**Affiliations:** Orthopaedic Technologies and Biomechanics Laboratory, Department of Mechanical Engineering, University of Victoria, 3800 Finnerty Road, Victoria BC V8P 5C2, Canada; School of Health Information Science, University of Victoria, 3800 Finnerty Road, Victoria BC V8P 5C2, Canada

**Author notes:** Corresponding author: Tess M. R. Carswell, PhD, Department of Mechanical Engineering, University of Victoria, 3800 Finnerty Road, Victoria BC V8P 5C2, Canada.

**Keywords:** Sex difference, Lower limb amputation, Rehabilitation, Prosthesis users, Patient-Oriented Research, Female lower limb prosthesis users

## Abstract

**Background:** Qualitative methods can facilitate understanding of the unique experiences of female lower limb prosthesis users (LLPUs) and inform sex-specific rehabilitation and prosthetic design. Given their underrepresentation in research, meaningful collaboration with female LLPUs is imperative to ensure efforts reflect their priorities.

**Objective:** Collaborate with female LLPUs to develop an interview guide with female-specific priorities and use it to investigate potential sex differences in needs of LLPUs.

**Study Design:** Qualitative research study.

**Methods:** Five female LLPUs were engaged to develop an interview guide, targeting aspects of a user’s prosthesis, and physical and psychosocial aspects of daily life. LLPUs were interviewed online and data were analyzed collaboratively using Codebook Thematic Analysis. Themes were extracted and assessed for sex differences.

**Results:** Interviews were conducted with 18 LLPUs (9-female, 9-male). Themes arose in four categories: 1-Prosthetic, 2-Physical, 3-Psychosocial, and 4-Financial Experiences, with sex differences observed in all but the financial category. Prosthetic themes revealed female-specific challenges with prosthetic fit, motivating future research into sex-specific prostheses. Physical themes expand understanding of female-specific ambulation challenges, presenting implications for sex-specific rehabilitation. Finally, psychosocial themes present new insight into sex-specific psychosocial experiences, including frustrations with clinicians and concerns regarding maternity and child-rearing.

**Conclusion:** Extracted themes demonstrated an array of sex-specific experiences, which can inform future research to address sex-specific rehabilitation and prosthetic design.

**Clinical Relevance:** This study identified sex-specific prosthetic, physical, and psychosocial challenges experienced by female LLPUs, underscoring the need for more sex-specific rehabilitation and prosthetic care. These findings can inform patient-centered clinical practice and guide future development of sex-specific prosthetic design and rehabilitation strategies.

## **1.** Introduction

Females are underrepresented in research on lower limb prosthesis users (LLPUs), including research on prosthetic design, satisfaction and biomechanical gait assessments.^1^ Despite females comprising ∼30-35% of the population of people with lower limb amputations,^2–5^ the proportion of female representation in a systematic review of prosthetic satisfaction was <14%,^6^ and in a review of lower limb prosthetic literature was ∼25%.^5^ Further, compared to males, females with lower limb amputations experience higher rates of mental health problems,^7^ greater challenges with ambulation and mobility,^8–12^ lower success with prosthetic fit,^12,13^ and greater prosthesis rejection rates and dissatisfaction.^4,12,14^

Despite growing evidence of sex and gender differences, further exploration is warranted to expand knowledge of female- and women-specific challenges and inform sex- and gender-specific rehabilitation and prosthetic design. Note, sex (e.g., female) and gender (e.g., women) are two distinct concepts. Sex is associated with biological and physiological features typically categorized as female/male and gender is a spectrum referring to psychological, social, and cultural factors influencing people’s roles, behaviors, and identities.^15^ While some have begun to explore sex and gender differences in prosthetic satisfaction, functional outcomes, and quality of life (QoL) aspects,^7^ many rely on generic measures or interview questions without consideration of female LLPU (fLLPU) or women LLPU (wLLPU) experiences to inform topics discussed. For example, Kuo et al.^16^ used the Orthotics and Prosthetics User’s Survey (OPUS),^17^ the Trinity Amputation and Prosthesis Experiences Scales (TAPES),^18^ and the Prosthetic Limb Users Survey of Mobility (PLUS-M) Short Form^19^ to study gender differences in satisfaction of prosthetic-related outcomes in veterans. Despite recruiting a substantial cohort of women (165/449 participants recruited), few gender differences were found, contrasting with the growing evidence that women report poorer overall outcomes.^7^ Kuo et al.^16^ noted these conflicting results may be due to their unique veteran-population benefiting from the extensive Veterans Affairs healthcare systems versus those available to the broader community. Further, Kuo et al.^16^ recognized that the generic measures used may not be able to adequately capture the unique experiences of wLLPUs, limiting their ability to accurately assess gender differences. Despite their popularity, generic measures like those used by Kuo et al.^16^ have been known to fail in capturing population-specific issues,^7^ potentially masking female- and women-specific challenges. Lehavot et al.^20^ conducted semi-structured interviews with veteran wLLPUs, asking about prosthetic experiences post-amputation. Some emergent themes echoed those of men, relating to importance of prostheses for mobility and QoL and others were women-specific such as challenges with clinician interactions, limited prosthetic options, and feelings of isolation.^20^ While these findings are highly relevant to uncover and understand challenges of fLLPUs and wLLPUs, this study only included veterans and also lacked consultation with fLLPUs or wLLPUs in study development. As described, veterans present a unique cohort with experiences not necessarily representative of the broader LLPU population. Further, without collaboration with fLLPUs or wLLPUs to guide research questions, it is unclear whether the questions asked and thus the topics broached by Lehavot et al.^20^ captured the full context and targeted highest priority topics for fLLPUs or wLLPUs.

In fact, few studies exploring sex and gender differences have meaningfully collaborated with fLLPUs and wLLPUs when conducting their research. When considering an underrepresented and underserved population, it is essential to collaborate with those of lived experience to ensure the research directions taken and the questions asked reflect the population’s unique priorities.^21,22^ Such efforts can be achieved by collaborating with fLLPUs and wLLPUs using Patient-Oriented Research (POR) methods to engage patients as partners in research.^21,22^ Incorporating POR and meaningfully including people with lived experience in the development and conduct of research has shown to improve its quality, transparency, and relevance.^23–25^

Therefore, we utilized POR to conduct a mixed-methods study in two phases, taking an explanatory sequential design.^26^ Phase-1 used POR to adapt the Prosthesis Evaluation Questionnaire (PEQ),^27^ and assess sex differences in LLPUs.^28^ The qualitative study described in this article is Phase-2, where POR was used to develop a semi-structured interview guide that better targets sex- and gender-related topics and key priorities of fLLPUs and wLLPUs. Using this interview guide, this study’s objective was to explore and expand understanding of sex and/or gender differences in the priorities and lived experience of LLPUs, focusing on prosthesis users with a unilateral transtibial (TTA) or transfemoral amputation (TFA). For simplicity, the cohort are referred to as LLPUs.

## 2. Methods

### 2.1 Study Design

This study took a qualitative descriptive approach,^29,30^ using individual semi-structured interviews to explore sex and gender differences in the priorities and lived experience of LLPUs. This approach was chosen as the aim was to stay close to the data, drawing on participant perceptions to accurately describe phenomena experienced.^29,30^ This work is reported according to the COREQ checklist.^31^ Ethics approval was obtained from the University of Victoria and the Vancouver Island Health Authority (H21-00778).

### 2.2 Participants, Setting, and Sampling

A subset of participants completing Phase-1^28^ were recruited for this qualitative work to expand upon sex and gender differences in perspectives and experiences of LLPUs and triangulate findings. Specifically, a convenience sample of participants from Phase-1^28^ were recruited to complete interviews until code saturation across the sample,^26,32^ while recruiting an equal number of female and male participants. Note, code saturation was identified when repetitive codes emerged during interview response analysis and new codes were not emerging even as additional interviews were completed.^26,32^

Participant eligibility included people with a unilateral TTA or TFA, aged 19-90 years with a Medicare Functional Classification Level (K-level) of 2 or higher, indicative of a limited community ambulator.^33^ Eligible participants could read and speak English and make decisions on their own behalf. Participants were ineligible if they had been medically diagnosed with severe depression, had their amputation for <1 year, were currently pregnant, had an osseointegrated prosthesis, or had a condition separate from their amputation severely affecting their ability to walk (e.g., stroke).

### 2.3 Research Team and Reflexivity

The research team comprised three researchers (TMRC, JWG, HM) and five fLLPUs (JC, LH, JN, DR, GT) referred to as Patient Partners (see acknowledgments). See Table 1 for contextual information about the team.

**Table 1.** Research team background and experience.

| Team Member | Sex | Gender | Occupation | Background |
|---|---|---|---|---|
| TMRC | Female | Woman | PhD Candidate<br>Mechanical Engineering | She has experience and training in POR and qualitative research, she has worked with LLPUs, and experience working with patients as participants. This study was undertaken as part of TMRC’s PhD. |
| HM | Female | Woman | Associate Professor<br>School of Health Information Science | She served on TMRC’s supervisory committee, and she has experience with POR and undertaking qualitative research in human factors and user experience. |
| JWG | Male | Man | Associate Professor<br>Mechanical Engineering | He was TMRC’s PhD supervisor, and he has experience in medical device design, working with patients as participants, and with POR. |
| JC | Female | Woman | Patient Partner<br>Research Team | She was a Patient Partner on the research team and has lived experience with a lower limb amputation and using a lower limb prosthesis. |
| LH | Female | Woman | Patient Partner<br>Research Team |  |
| JN | Female | Woman | Patient Partner<br>Research Team |  |
| DR | Female | Woman | Patient Partner<br>Research Team |  |
| GT | Female | Woman | Patient Partner<br>Research Team |  |

Abbreviations: POR, Patient-Oriented Research; LLPUs, lower limb prosthesis users.

Due to the small population of LLPUs where the study was based (i.e., Victoria, BC), some participants were known to the team in a research context (e.g., participated in past research). All participants had knowledge of the research purpose since background information was included in the consent form.

The research team practiced reflexivity,^34^ engaging in conversations through different stages of the research about their subjectivity, lived experience, and personal background and how these may shape the work conducted and their perspectives on it. The team also engaged in other common reflexive practices, including transcript verification.^35,36^ Finally, the POR approach taken and direct involvement of people with lived experience (i.e., Patient Partners) in the creation of research design engrained reflexivity within this work.^37^ Iteratively consulting and collaborating with the Partners helped to ensure actions taken and decisions made through design, data collection, and analysis were shaped communally.^37^

### 2.4 Interview Guide and Data Collection Procedure

To ensure information garnered from interviews was relevant to fLLPU and wLLPU priorities and to the study’s purpose, questions were derived in collaboration with the research team (Table 2). Team meetings informed topics covered in the semi-structured interview guide, including perspectives and experiences with lower limb loss, and unmet needs and priorities for prostheses and QoL outcomes. Derived questions resembled those of the adapted questionnaire instrument from the Phase-1,^28^ as mentioned taking an explanatory sequential design to expand understanding of sex and/or gender differences in experiences of LLPUs derived in Phase-1 and triangulate findings between phases. Probing questions were asked when necessary, encouraging participants to elaborate and expand on their responses to obtain more detailed information. Once the interview guide was developed, a pilot interview was conducted with two Patient Partners (JC, LH). Note, while Partners informed topics covered in the interview guide, they were not necessarily involved in deciding order, phrasing, or overall study procedures. Therefore, the pilot interviews provided relevant feedback on these aspects.

**Table 2.** Semi-structured interview guide.

| Question |
|---|
| ▪ Do you think your needs are met with your current prosthesis? Please explain. |
| - If ‘no’ – How could your prosthesis be improved to meet those? |
| ▪ Do you think your priorities are met with your current prosthesis? Please explain. |
| - If ‘no’ – How could your prosthesis be improved to meet those? |
| - Please rank your top three priorities and explain this ranking. |
| ▪ Are you satisfied with your mobility? Please explain |
| - If ‘no’ – What is something that could improve that? |
| ▪ Are you satisfied with the support you have received from medical professionals since your amputation for your physical health? Please explain. |
| - If ‘no’ – How do you think that support could be improved? |
| ▪ Are you satisfied with the support you have received from medical professionals since your amputation for your mental health? Please explain. |
| - If ‘no’ – How do you think that support could be improved? |
| ▪ Has your amputation affected your interest in having children? |
| - If so, how? |
| - Has it affected your parenting and if so, how? |
| ▪ Do you feel as if your self-esteem and/or confidence has been affected since your amputation? |
| - If so, how? |
| ▪ Do you feel as if your masculinity/femininity has been affected since your amputation? |
| - If so, how? |
| ▪ Has your independence been affected since your amputation? |
| - If so, how? |
| ▪ Please describe your ideal prosthesis in what it would look like, feel like, and how it would behave. |

All interviews were conducted by the lead researcher (TMRC) over Zoom (December 2021-April 2023). A second researcher (JWG) was present for the first two interviews. Interviews were conducted in the presence of only the participant (i.e., interviewee) and interviewer(s), with participants ensuring they were in a quiet, private space. Prior to recording, the interviewer confirmed participants had read and understood the consent form and following this, participants gave verbal consent. Once the audio and video recording started, participants re-iterated verbal consent for documentation purposes. Next, the lead interviewer followed the semi-structured interview guide (Table 2). Before concluding, participants were informed that they would have the opportunity to review and verify their transcripts.^36^ No field notes were taken during or after interviews and only one interview was conducted per participant. Session duration ranged from 31-93min (average: 61min).

### 2.5 Transcript Verification and Data Analyses

Demographic data were collected in Phase-1^28^ and interview participant responses, including demographic data, were analyzed between sexes and genders using Mann-Whitney U or independent samples *t*-tests as appropriate. Statistical significance was set to *p*<0.05. Statistical analyses were performed in SPSS v27.0 (IBM Corp., Armonk, NY).

Interviews were transcribed verbatim (aided by Zoom transcription software), reviewed and anonymized by the lead researcher (TMRC). Participants verified transcript accuracy within two weeks of their interview.^36^

Interview data were analyzed using Codebook Thematic Analysis with a template analysis appoach,^38–40^ and collaborative efforts guided by the DEPICT method (Table).^41^ Coupling Codebook Thematic Analysis with the DEPICT method enables a practical approach to collaborative qualitative analysis amongst the research team, to meaningfully engage Patient Partners, enhance rigour of analysis and relevance of results through inclusion of stakeholders with diverse lived experiences.^41^ One researcher (TMRC) and two Patient Partners (DR, GT) were involved in all steps of data analysis (referred to as the analysis cohort) while the remainder of the research team devoted efforts in later stages to collaboratively analyzing, translating, and resolving uncertainty around findings. The process followed by the analysis cohort for each step of the DEPICT method is outlined in Table. See Supplemental Digital Content 1 (SDC-1) for the finalized codebook.

Note, *a priori* thresholds were established to help guide analysis (Table 3). Specifically, a 60% (n=6/9) threshold was considered as a majority metric, to help guide consideration for a “commonly” reported theme.^42^ As well, one sex reporting something >30% (n=3/9) more than another provided a guide for sex-specific prioritization. While not necessarily common in qualitative research, others have used *a priori* thresholds to help guide thematic review.^28,43^ As well, when taking raw qualitative data and contextualizing it into grouped “common” themes, researchers are establishing some basis for what is “common”.^42^ In this way, utilizing thresholds can help to establish said patterns and generate starting hypotheses.^42^ Doing so, we can help avoid pitfalls of overweighing one participant’s particular account over another due to our own biases, how verbose one account is over another, etc…^42^ However, prevalence alone did not dictate thematic importance, and these established thresholds were not necessarily deciding factors for themes considered. Depth and richness of insight, nuance provided, along with prevalence were considerations for thematic importance and distinguishers for sex-specific vs sex-agnostic themes.

**Table 3.**
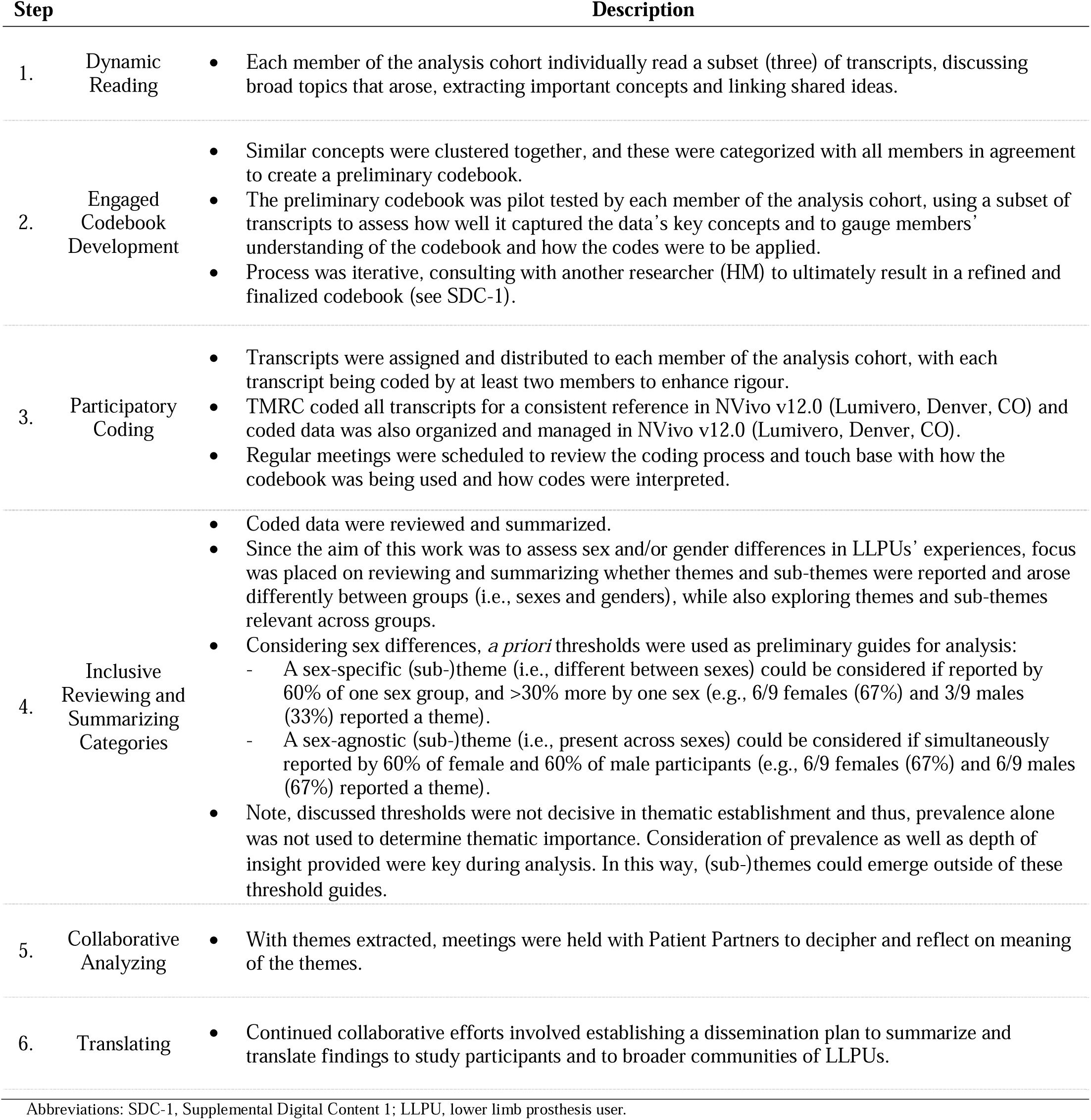
Process followed for each step of the DEPICT method.

| Step |  | Description |
|---|---|---|
| 1. | Dynamic Reading | • Each member of the analysis cohort individually read a subset (three) of transcripts, discussing broad topics that arose, extracting important concepts and linking shared ideas. |
| 2. | Engaged Codebook Development | • Similar concepts were clustered together, and these were categorized with all members in agreement to create a preliminary codebook.<br>• The preliminary codebook was pilot tested by each member of the analysis cohort, using a subset of transcripts to assess how well it captured the data’s key concepts and to gauge members’ understanding of the codebook and how the codes were to be applied.<br>• Process was iterative, consulting with another researcher (HM) to ultimately result in a refined and finalized codebook (see SDC-1). |
| 3. | Participatory Coding | • Transcripts were assigned and distributed to each member of the analysis cohort, with each transcript being coded by at least two members to enhance rigour.<br>• TMRC coded all transcripts for a consistent reference in NVivo v12.0 (Lumivero, Denver, CO) and coded data was also organized and managed in NVivo v12.0 (Lumivero, Denver, CO).<br>• Regular meetings were scheduled to review the coding process and touch base with how the codebook was being used and how codes were interpreted. |
| 4. | Inclusive Reviewing and Summarizing Categories | • Coded data were reviewed and summarized.<br>• Since the aim of this work was to assess sex and/or gender differences in LLPUs’ experiences, focus was placed on reviewing and summarizing whether themes and sub-themes were reported and arose differently between groups (i.e., sexes and genders), while also exploring themes and sub-themes relevant across groups.<br>• Considering sex differences, *a priori* thresholds were used as preliminary guides for analysis:<br>- A sex-specific (sub-)theme (i.e., different between sexes) could be considered if reported by 60% of one sex group, and >30% more by one sex (e.g., 6/9 females (67%) and 3/9 males (33%) reported a theme).<br>- A sex-agnostic (sub-)theme (i.e., present across sexes) could be considered if simultaneously reported by 60% of female and 60% of male participants (e.g., 6/9 females (67%) and 6/9 males (67%) reported a theme).<br>• Note, discussed thresholds were not decisive in thematic establishment and thus, prevalence alone was not used to determine thematic importance. Consideration of prevalence as well as depth of insight provided were key during analysis. In this way, (sub-)themes could emerge outside of these threshold guides. |
| 5. | Collaborative Analyzing | • With themes extracted, meetings were held with Patient Partners to decipher and reflect on meaning of the themes. |
| 6. | Translating | • Continued collaborative efforts involved establishing a dissemination plan to summarize and translate findings to study participants and to broader communities of LLPUs. |

Abbreviations: SDC-1, Supplemental Digital Content 1; LLPU, lower limb prosthesis user.

## 3. Results

### 3.1 Participant Characteristics

Of the 28 participants from Phase-1,^28^ a total of 20 interviews were completed before reaching code saturation across the full sample.^26^ No participants were lost to attrition, however, following completion of both phases, two participants were excluded due to ineligibility. Data from these two participants was excluded in both phases. Responses from the remaining 18 (9-females, 9-males) eligible participants from Phase-2 were analyzed (Table 4). For individual participant demographics, see SDC-2.

**Table 4.** Summary of participant demographics total and stratified by sex.

| **Demographic Parameter** | **Total** | **Female** | **Male** | ***p*-value** |
|---|---|---|---|---|
|  | Mean (SD) / n (% of 18) | Mean (SD) / n (% of 9) | Mean (SD) / n (% of 9) |  |
| Age (years) | 58.4 (19.4) | 53.6 (24.7) | 63.3 (11.7) | 0.306 |
| BMI (kg/m<sup>2</sup>) | 26.7 (4.73) | 25.4 (5.08) | 28.0 (4.26) | 0.262 |
| Gender Identity<sup>*</sup> |  |  |  | - |
| Woman | 9 (50.0) | 9 (100) | 0 (0.00) | - |
| Man | 9 (50.0) | 0 (0.00) | 9 (100) | - |
| Self-Described | 0 (0.00) | 0 (0.00) | 0 (0.00) | - |
| Amputation Level |  |  |  | 0.113 |
| TTA | 14 (77.8) | 9 (100) | 5 (55.6) | - |
| TFA | 4 (22.2) | 0 (0.00) | 4 (44.4) | - |
| Time Since Amputation (years) |  |  |  | 0.730 |
| 1-2 | 5 (27.8) | 3 (33.3) | 2 (22.2) | - |
| 3-5 | 3 (16.7) | 2 (22.2) | 1 (11.1) | - |
| 6-10 | 2 (11.1) | 0 (0.00) | 2 (22.2) | - |
| >10 | 8 (44.4) | 4 (44.4) | 4 (44.4) | - |
| Time with Current Prosthesis (years) |  |  |  | 0.222 |
| <1 | 7 (38.9) | 5 (55.6) | 2 (22.2) | - |
| 1-2 | 3 (16.7) | 1 (11.1) | 2 (22.2) | - |
| 3-5 | 7 (38.9) | 3 (33.3) | 4 (44.4) | - |
| >5 | 1 (5.56) | 0 (0.00) | 1 (11.1) | - |
| Amputation Pathology |  |  |  | 0.931 |
| Vascular | 5 (27.8) | 3 (33.3) | 2 (22.2) | - |
| Trauma | 8 (44.4) | 3 (33.3) | 5 (55.5) | - |
| Cancer | 1 (5.56) | 1 (11.1) | 0 (0.00) | - |
| Other | 4 (22.2) | 2 (22.2) | 2 (22.2) | - |
| Geographic Location |  |  |  | 0.387 |
| BC, Canada | 11 (61.1) | 7 (77.8) | 4 (44.4) | - |
| AB, Canada | 4 (22.2) | 0 (0.00) | 4 (44.4) | - |
| ON, Canada | 1 (5.56) | 1 (11.1) | 0 (0.00) | - |
| Undisclosed | 2 (11.1) | 1 (11.1) | 1 (11.1) | - |

Abbreviations: SD, standard deviation; BMI, body mass index; TTA, transtibial amputation; TFA, transfemoral amputation; BC, British Columbia; AB, Alberta; ON, Ontario.
Note\*: Gender identity is presented as three discrete options in this table for simplicity. However, this question regarding gender identity was posed as an open-ended question, left for participants to fill out as they saw fit.

As the cohort comprised only of cisgender people (i.e., sex assigned at birth aligned with gender identity), there were not enough gender-diverse perspectives to distinguish between sex versus gender differences. Therefore, stratifications and analyses were performed based on sex only.

There were no statistically significant differences in demographics between female and male cohorts (Table 4).

### 3.2 Resulting Themes and Sub-Themes

Themes were grouped into four categories, 1-Prosthetic, 2-Physical, 3-Psychosocial, and 4-Financial. Within each category, some themes and sub-themes were sex-agnostic whereas others were sex-specific. Figure 1 summarizes the hierarchical themes and sub-themes emerging, with female sex-specific themes in pink, male sex-specific themes in blue, and sex-agnostic themes in orange.

**Figure 1.**
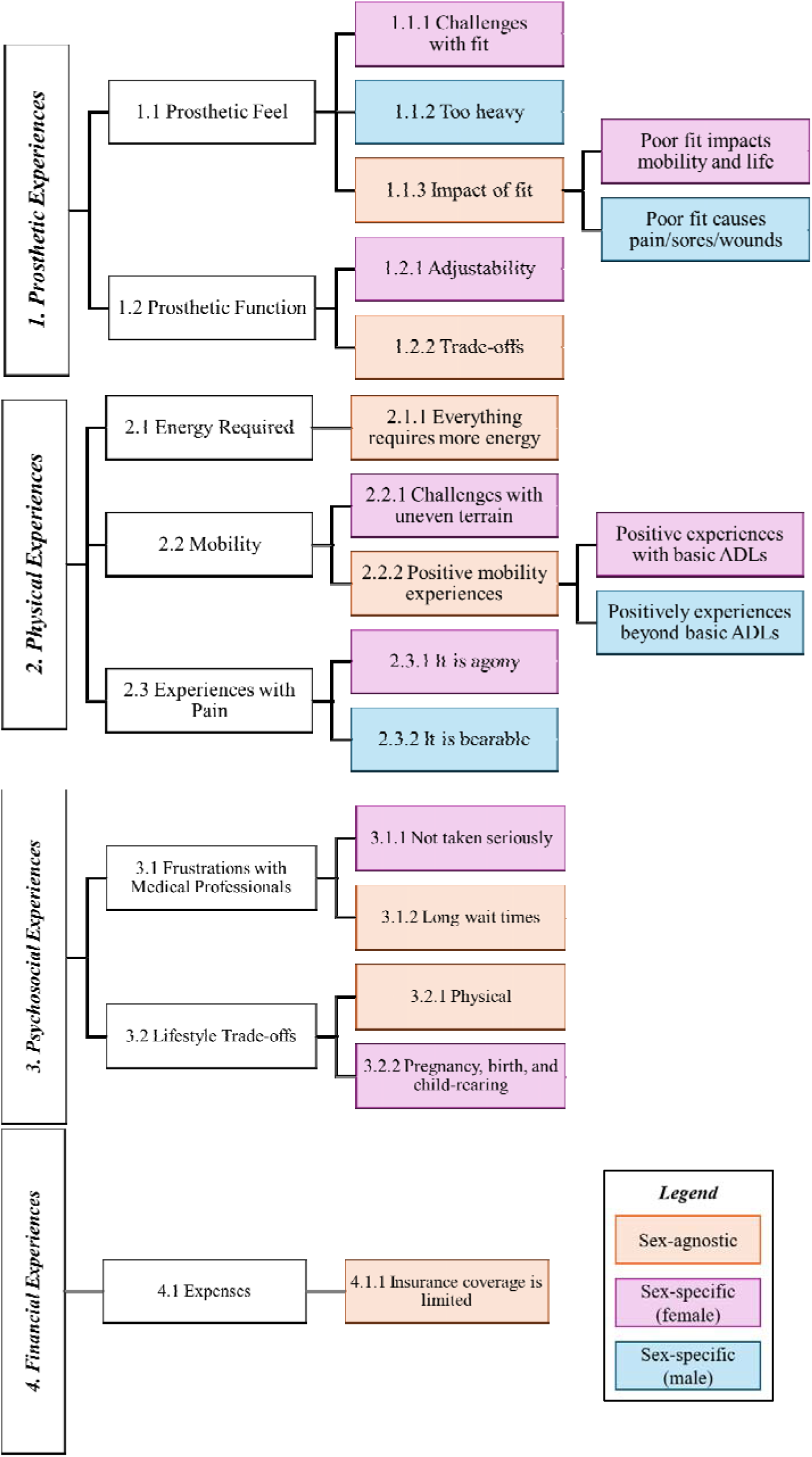
Summary of themes emerging from LLPUs open-ended question responses. (Abbreviations: ADLs, activities of daily living)

Table 5 summarizes the sub-themes emerging from these categories, whether they were sex-specific or sex-agnostic. Note, while some (sub-)themes were sex-agnostic, reports of the (sub-)theme might have been sex-specific. Further, a sex-specific (sub-)theme does not necessarily mean that only one sex reported it, both sexes may have reported it. However, it was distinguished as sex-specific through consideration of prevalence as well as depth of insight and nuance provided by one sex over another (Table 3).

**Table 5.** Themes emerging from open-ended question responses.

| ***Category***<br>**Themes**<br>**Sub-Theme** | **Theme-Type** | **Description** | **Emblematic Quote(s)** |
|---|---|---|---|
| *1. Prosthetic Experiences* |  |  |  |
| 1.1 Prosthetic Feel |  |  |  |
| 1.1.1 Challenges with fit | Sex-specific (Females) | F: Muscle loss and shape changes negatively impact prosthesis fit | F002: *“…it has a lot to do with the shape of my limb… due to the loss of a lot of the muscle around it- it does mean that there are spots in it that aren't touching the liner properly and I believe it, therefore, means that it isn’t interacting with the socket properly.”*<br>F003: *“… although I think in the past, you know, maybe the hormonal every month, like carrying a bit more fluid in your body as a woman would change the fit of a prosthetic.”* |
| 1.1.2 Too heavy | Sex-specific (Males) | M: Prostheses are too heavy | M009: *“And lightweight, the lighter the better. If [prostheses] could weigh one ounce, you know, like one gram that would be great, that would be a perfect prosthetic to have that you didn't even know was there it was so light.”*<br>M013: *“I think, you never get used to the weight of it, you know, like they say it's the same weight basically as what the leg was, but man oh man that thing feels heavy.”* |
| 1.1.3 Impact of fit | Sex-agnostic | F/M: Poor fit impacts mobility and life | F003: *“Comfort is definitely first and foremost on my mind when I put on my leg in the morning. Without comfort, I don't have mobility like I want or need.”* |
|  |  | F/M: Poor fit causes pain/sores/wounds | M008: *“So it makes your prosthetic either looser or tighter as the day goes on, and that can cause fatigue in your residual limb. So by the end of the day, you know, it can be kind of sore and cause phantom pain, or, you know, real pain in your, in your short limb.”* |
| 1.2 Prosthetic Function |  |  |  |
| 1.2.1 Adjustability | Sex-specific (Females) | F: Ankle mobility is a priority | F002: *“Feet with more spring and a natural ankle motion, greatly improve mobility.”*<br>F010: *“Well, it would be great if I had an ankle that was like, and they do make them, they make like the really fancy ankles that can adjust to inclines and stuff.”* |
| 1.2.2 Trade-offs | Sex-agnostic | F/M: With prostheses, you will always have to sacrifice one thing to get another | F003: *“I’d like the option of having that adjustable heel. But again, I have to figure out whether I’m having a leg that's gonna help me with my mobility and the physical activities overall or whether I want something that's a little bit more geared toward aesthetics. I don't get coverage for both. So I have to be very conscious of making smart choices with what will provide the best long-term mobility for all activities.”* |
| *2. Physical Experiences* |  |  |  |
| 2.1 Energy Required |  |  |  |
| 2.1.1 Everything requires more energy | Sex-agnostic | F/M: Everything with an amputation requires more energy | F003: *“It takes a lot of energy, just no matter what you're doing, as an amputee. It just takes more energy and a lot more thought process, even when walking.”*<br>M007: *“Basically standing, pedaling, moving… like basically even just standing burns energy.”* |
| 2.2 Mobility |  |  |  |
| 2.2.1 Challenges with uneven terrain | Sex-specific (Females) | F: Challenges experienced on non-flat terrain | F010: *“[Walking] gets harder of course, like on less even surfaces. I find mobility on really steep inclines is difficult, particularly downhill but both difficult.”*<br>F012: *“When there's steps and uneven kinds of things I have to, I have to do it very consciously.”*<br>F014: *“You have to pay more attention with how you're walking. Otherwise, walk on like flat pavement, no problem. But uneven [terrain], little bit more chance to have an issue with your gait.”* |
| 2.2.2 Positive mobility experiences | Sex-agnostic | F: Positive experiences are limited to basic activities of daily living (ADLs) | F010: *“I’m happy with kind of the more base-level mobility… like yeah getting up and down is fine getting yeah walking around is fine.”* |
|  |  | M: Positive experiences extend beyond basic ADLs | M015: *“Honestly, it's great, like bike riding, I can go golfing, it doesn't affect me when I go into the mall, or you know I’m able to walk great distances like luckily enough. I’ll put it in stepwise because of the fit-bits and things, and we were doing like 15,000 steps a day.”* |
| 2.3 Experiences with Pain |  |  |  |
| 2.3.1 It is agony | Sex-specific (Males) | M: Reports of pain are severe and intense | M006: *“I went to a hockey game with my kids, and I sat there in agony because I was so uncomfortable.”* |
| 2.3.2 It is bearable | Sex-specific (Females) | F: Reports of pain are manageable | F012: *“There's always some kind of pain, but it's manageable and I kind of can get used to it.”* |
| *3. Psychosocial Experiences* |  |  |  |
| 3.1 Frustrations with Medical Professionals |  |  |  |
| 3.1.1 Not taken seriously | Sex-specific (Females) | F: Not being taken seriously or listened to my medical professionals | F002: *“I would like my prosthetist, doctor, and/or physiatrist to help me navigate this issue. They never seem to take it seriously. That would be great. When it comes to the shape of the prosthetic, my prosthetist never asks what I want or need. I just need my practitioners to listen.”* |
| 3.1.2 Long wait times | Sex-agnostic | M/F: Healthcare is slow, doctors are busy, so you need to self-advocate and do things yourself | M006: *“The medical profession is pretty much a joke in this country, and you have to take responsibility for your own physical well-being, mental well-being and all that.”* |
| 3.2 Life-style Changes and Choices |  |  |  |
| 3.2.1 Physical activity trade-offs | Sex-agnostic | M/F: Physical activities are limited now | M011: *“I used to do a lot of hiking, and a lot of tree climbing, and all that kind of stuff and I can't do that, and I don't do that and I don't try anymore. So I understand my restrictions, and I’m learning to live with that.”* |
| 3.2.2 Pregnancy, birth, and child-rearing | Sex-specific (Females) | F: Uncertainty and new concerns around having kids | F002: *“It has made it a lot harder to want children, imagining myself as an amputee-mother.”*<br>F003: *“The only real worry I had was keeping up with my kids. How would I physically keep up with them?”*<br>F010: *“It has made me more stressed about it, and it has, like adjusted our timeline of when we would start trying for kids. I do worry about it.”*<br>F016: *“I think I’m just unsure if my body can handle it... the giving birth part.”* |
| *4. Financial Experiences* |  |  |  |
| 4.1 Expenses |  |  |  |
| 4.1.1 Insurance coverage is limited | Sex-agnostic | F/M: Insurance only covers very basic prostheses and limits even basic activities | F003: *“We get covered for basic mobility. We don't get covered for individual consideration and individual needs or priorities regarding what mobility looks like for each one of us.”*<br>M006: *“I was recently denied a running prosthesis because physical activity is not seen as a “necessity”… So in my mind, they’re choosing to ignore my physical and mental well-being, which there's a whole lot of documentation about the benefits and the necessity of movement, etc.”* |

Abbreviations: F, female; M, male; ADLs, activities of daily living.

#### 3.2.1 Prosthetic Experiences

Themes relating to Prosthetic Feel (1.1) and Prosthetic Function (1.2) emerged, relaying experiences of how the prosthesis feels on the body and how the prosthesis functions, respectively. Within ‘Prosthetic Feel’, three sub-themes emerged: Challenges with fit, Too heavy, and Impact of fit. Within ‘Prosthetic Function,’ two sub-themes emerged: Adjustability, and Trade-offs.

##### Challenges with fit (1.1.1) – Female-specific

Both female and male participants discussed challenges with prosthetic fit. However, male reports (n=2) were unprolific and did not provide rich substance into their actual challenges with fit. Conversely, all female participants (n=9) expressed such challenges, delving into detail and intention, distinguishing this as a sex-specific sub-theme. Some female participants relayed that muscle loss was the culprit, negatively impacting fit of their prosthesis.

> *“…it has a lot to do with the shape of my limb… due to the loss of a lot of the muscle around it-it does mean that there are spots in it that aren’t touching the liner properly and I believe it, therefore, means that it isn’t interacting with the socket properly.” (F002)*

Other females reported that fluid volume fluctuations resulting from their menstrual cycle or general monthly hormonal changes could be causing challenges with prosthetic fit.

> *“… although I think in the past, you know, maybe the hormonal every month, like carrying a bit more fluid in your body as a woman would change the fit of a prosthetic.” (F003)*

##### Too heavy (1.1.2) – Male-specific

Both females and males expressed concerns about the weight of their prosthesis. However, this was not as prioritized a topic for female participants (n=3) as it was for males (n=7), with females emphasizing other topics around Prosthetic Feel (e.g., *Challenges with fit - 1.1.1*), distinguishing this as a sex-specific sub-theme. Male participants discussed their satisfaction within Prosthetic Feel around prosthetic weight, idealizing a lightweight device.

> *“And lightweight, the lighter the better. If [prostheses] could weigh one ounce, you know, like one gram that would be great, that would be a perfect prosthetic to have that you didn’t even know was there it was so light.” (M009)*

Similarly, male participants criticized how heavy their prosthesis felt to them, even if this was more so their perception of the weight.

> *“I think, you never get used to the weight of it, you know, like they say it’s the same weight basically as what the leg was, but man oh man that thing feels heavy.” (M013)*

##### Impact of fit (1.1.3) – Sex-agnostic

Across all participants (n=12), there was prolific discussion around how prosthesis fit impacts other aspects of their life and health, distinguishing this as a sex-agnostic sub-theme. Several participants discussed how fit impacts health of their residual limb, causing discomfort, sores or general fatigue.

> *“So it makes your prosthetic either looser or tighter as the day goes on, and that can cause fatigue in your residual limb. So by the end of the day, you know, it can be kind of sore and cause phantom pain, or, you know, real pain in your, in your short limb.”(M008)*

Others focused more broadly on how fit impacts their daily life and mobility as opposed to specific physical impacts.

> *“Prosthetic fit is a huge part of that mobility and comfort is what enables me to do what I do everyday. This comfort is definitely first and foremost on my mind when I put on my leg in the morning. Without comfort, I don’t have mobility like I want or need.” (F003)*

##### Adjustability (1.2.1) – Female-specific

The great majority of female participants (n=8) discussed prosthetic function relating to adjustability at the ankle. While some male participants reported similar aspects of ankle adjustability (n=4), these accounts were more transient than the female perspectives, distinguishing this as a sex-specific sub-theme. Female participants reported ankle adjustability as a priority since it would allow for improved mobility and more natural biomechanics.

> *“Feet with more spring and a natural ankle motion, greatly improve mobility.” (F002)*

Other female participants shared how improved ankle range of motion (ROM) could aid with different types of terrain, including incline.

> *“Well, it would be great if I had an ankle that was like, and they do make them, they make like the really fancy ankles that can adjust to inclines and stuff. Helpful for mobility. But they’re very expensive and less good at other things. So I find when I’m choosing a prosthesis, if I’m choosing one that I’m going to be wearing most of the time, I want it to kind of be more of a Jack-of-all-trades than like a king-of-one-trade.” (F010)*

##### Trade-offs (1.2.2) – Sex-agnostic

Across participants (n=13), there was significant discussion regarding trade-offs for prosthetic function, marking this as a sex-agnostic sub-theme. When referring to satisfaction or frustrations with their prostheses, participants would often reference the give and take, and needing to sacrifice one aspect for another.

> *“It is heavy. Like, the knee I got is quite heavy. But, it also does a lot of work for me. Like I have this mechanical one here, it’s light as a feather but I have to make it bend, you know. And I gotta swing my hip forward to lock it back in. So it’s actually a lot more work, where this one does it for me. No, it’s heavy but it pulls its weight.” (M004)*

Another cited trade-off related to mobility versus aesthetics, and expense and insurance coverage playing a role with these decisions.

> *“I’d like the option of having that adjustable heel. But again, I have to figure out whether I’m having a leg that’s gonna help me with my mobility and the physical activities overall or whether I want something that’s a little bit more geared toward aesthetics. I don’t get coverage for both. So I have to be very conscious of making smart choices with what will provide the best long-term mobility for all activities.” (F003)*

As well, the earlier quote by F010 spoke to female participant’s prioritizing ankle adjustability. However, this concept of trade-offs also emerges within their statement. F010 shared this need to balance prosthetic function with cost and looking for some sort of all-in-one option.

#### 3.2.2 Physical Experiences

Themes relating to Energy Required (2.1), Mobility (2.2), and Experiences with Pain (2.3) emerged, relaying physical experiences of vitality, movement, and pain (respectively) when living with an amputation. Within ‘Energy Required’, one sub-theme arose: Everything requires more energy. Within ‘Mobility’, two sub-themes arose: Challenges with uneven terrain, and Positive mobility experiences. Within ‘Experiences with Pain’, two sub-themes arose: It is agony, and It is bearable.

##### Everything requires more energy (2.1.1) – Sex-agnostic

A sex-agnostic sub-theme emerged, with participants (n=14) sharing how everything requires more energy when you are living with limb loss. This expenditure related to both physical and mental loads.

> *“It takes a lot of energy, just no matter what you’re doing, as an amputee. It just takes more energy and a lot more thought process, even when walking.” (F003)*

##### Challenges with uneven terrain (2.2.1) – Female-specific

When discussing mobility, all female participants (n=9) reported challenges traversing uneven terrain, something not emphatically discussed in the male cohort, distinguishing this as a female-specific sub-theme. Females emphasized having challenges with their movement, specifically on hills.

> *“[Walking] gets harder of course, like on less even surfaces. I find mobility on really steep inclines is difficult, particularly downhill but both difficult.” (F010)*

As well, female participants reported having difficulty not just on steep hills but broadly on non-flat terrain.

> *“You have to pay more attention with how you’re walking. Otherwise, walk on like flat pavement, no problem. But uneven [terrain], little bit more chance to have an issue with your gait.” (F014)*

##### Positive mobility experiences (2.2.2) – Sex-agnostic but reported differently

Both female and male participants discussed having positive experiences with mobility. However, how these positive experiences were characterized differed by sex. When females discussed their positive experiences, these were limited to basic activities of daily living (ADLs) (n=5).

> *“I’m happy with kind of the more base-level mobility… like yeah getting up and down is fine getting yeah walking around is fine.”(F010)*

Conversely, male participants reported on mobility experiences vastly extending beyond basic ADLs, including more athletic endeavors (n=8).

> *“Honestly, it’s great, like bike riding, I can go golfing, it doesn’t affect me when I go into the mall, or you know I’m able to walk great distances like luckily enough. I’ll put it in stepwise because of the fit-bits and things, and we were doing like 15,000 steps a day.” (M015)*

##### It is agony (2.3.1) – Male-specific

All participants reported experiences of pain (n=18). However, a sub-theme within male experiences saw participants (n=5) providing accounts that were intense and severe.

> *“I went to a hockey game with my kids, and I sat there in agony because I was so uncomfortable. I was like, is this normal? Like… is this kind of the way it goes? Or should I be able to sit and have a dinner with my wife, or whatever, and not be fidgeting and just dying to take my leg off?” (M006)*

##### It is bearable (2.3.2) – Female-specific

Conversely, female participants reported regularly having experiences with pain but a sub-theme within their accounts reported pain as manageable (n=6).

> *“There’s always some kind of pain, but it’s manageable and I kind of can get used to it.” (F012)*

#### 3.2.3 Psychosocial Experiences

Themes arose relating to Frustrations with Medical Professionals (3.1) and Life-style Changes and Choices (3.2) emerged, with participants sharing their challenges and dissatisfaction with the healthcare system and clinicians, and relaying the changes and choices they were having to make due to their amputation. Within ‘Frustrations with Medical Professionals’, two sub-themes emerged: Not taken seriously, and Long wait times. Within ‘Life-style Changes and Choice’, two sub-themes emerged: Physical activity trade-offs, and Pregnancy, birth, and child-rearing.

##### Not taken seriously (3.1.1) – Female-specific

When discussing interactions and satisfaction with medical professionals, all female participants (n=9) reported not feeling listened to and not being taken seriously. This theme was not reported by males (n=0), distinguishing it as a female-specific theme. Female participants shared that their priorities were not taken seriously.

> *“I have had to be creative in finding ways to help deal with the sweat and lack of suction without irritating the skin but nothing is perfect. I would like my prosthetist, doctor, and/or physiatrist to help me navigate this issue. They never seem to take it seriously. That would be great. When it comes to the shape of the prosthetic, my prosthetist never asks what I want or need. I just need my practitioners to listen.” (F002)*

Female participants also emphasized needing to self-advocate to counteract not being listened to or taken seriously.

> *“I’ve been to my doctor twice, I’ve been to the surgeon once, and none of them have looked at my leg. It’s just, we talk and they don’t listen. And then they say “well take these pills, it should go away”. No, no. I don’t want that. I guess… the more you complain, the more you get action.” (F001)*

##### Long wait times (3.1.2) – Sex-agnostic

Across all participants (n=18), a reoccurring sub-theme discussed long wait times within the healthcare system, improper access to medical professionals and often feeling as if clinicians were too busy to give proper care. While in the previous sex-specific sub-theme, females reported self-advocation to counteract not being listened to or taken seriously, in this sex-agnostic sub-theme, both female and male participants reported needing to self-advocate to counteract long wait times and busy clinicians.

> *“The medical profession is pretty much a joke in this country. Not one has time and you have to take responsibility for your own physical well-being, mental well-being and all that.” (M006)*

Similar sentiments of slipping through the healthcare system’s cracks were echoed by other participants.

> *“And I didn’t really want to sign any of the paperwork, I was scared I was gonna get lost in the system. You know, because everything was just going and begin to the rehab, we had to chase them down. You know all that stuff kind of going. We didn’t know who to call, like there’s no “Hey, you just lost a leg. Here, here’s a book for amputees.” (M004)*

##### Physical activity trade-offs (3.2.1) – Sex-agnostic

Across the majority of female (n=7) and male (n=8) participants, a sex-agnostic theme arose with participants sharing that they have had to limit the physical activities they do or completely stop some of the activities they used to do because of their amputation.

> *“I used to do a lot of hiking, and a lot of tree climbing, and all that kind of stuff and I can’t do that, and I don’t do that and I don’t try anymore. So I understand my restrictions, and I’m learning to live with that quite easily.” (M011)*

##### Pregnancy, birth, and child-rearing (3.2.2) – Female-specific

The majority of female participants (n=5) shared newfound concerns and overall uncertainty around having children. These points were not reported by male participants (n=0), distinguishing this as a female-specific sub-theme. Some female participants shared concerns around physical aspects of child-rearing.

> *“The real worry I had was keeping up with my kids. How would I physically keep up with them? I’ve got three boys, and I just adapted with the first one. And when I found out I was pregnant with the second one, it really worried me. Yeah, obviously, because young kids are super active and their safety came into question too, because what if they bolt in the street? Can I catch up to them? You know, those were the kind of concerns, for sure. I think those are normal worries for all parents on some level… but as an amputee, I think you definitely consider these little things more.” (F003)*

Other female participants shared concerns around pregnancy, giving birth, and the toll on their body.

> *“Yeah, I think I um… it’s interesting because, like I think I’m just unsure if my body can handle it… The giving birth part, yeah, probably not. I think I would be okay with adoption maybe, that would be fine. But like that’s not yeah, it’s not as much on my body.” (F017)*

Concerns over balancing the expenses of a child on top of regular medical expenses were raised, as well as the physical toll and having enough energy for it all.

> *“…it has made it a lot harder to want children, imagining myself as an amputee-mother. My husband’s had to help me out so much as it is, and the thought of him having to take on a baby as well is a lot. If anything goes wrong with me, especially since complications with my amputation seem to continuously arise, it scares me that he would have to deal with so much-work and finances included. Getting up in the middle of the night, trying to balance the cost of being an amputee and having a baby, imagining myself keeping up with, or having enough energy to care for a child, dealing with the extra weight during and after pregnancy…that kind of stuff is difficult.”(F002)*

#### 3.2.4 Financial Experiences

A theme of Expenses (4.1) emerged, with participants discussing the cost of living with an amputation and being a prosthesis user. Within ‘Expenses’, one sub-theme arose: Insurance coverage is limited.

##### Insurance coverage is limited (4.1.1) – Sex-agnostic

The majority of female (n=8) and male participants (n=6) discussed the vast limitations of their insurance and that coverage is extremely basic and out of touch with the needs of LLPUs.

> *“I’m lucky that I have extended health insurance as well but often that doesn’t even cover needs. Extended health coverage lacks understanding of the realities of amputee life and how certain components are necessary to fit a prosthetic leg. For example, they will not cover the liners that I need to actually don/wear my leg. Pharmacare will cover two a year but when you alternate two every day, all day, all year, it’s similar to asking a person to wear the same pair of socks for a year.” (F003)*

Other participants shared that their limited coverage impacts mobility needs, with conflicting perceptions on what should be considered basic mobility and what within this is a basic necessity.

> *“I was recently denied a running prosthesis because physical activity is not seen as a “necessity”… So in my mind, they’re choosing to ignore my physical and mental well-being, which there’s a whole lot of documentation about the benefits and the necessity of movement, etc.” (M006)*

## 4. Discussion

This study utilized POR to develop an interview guide that better targets key priorities of fLLPUs and wLLPUs and explored understanding of sex and gender differences in the lived experience of LLPUs. Themes arose from interview responses and were organized into four categories (Table 5). Within these categories, various (sub-)themes arose differently between sexes except for those within financial experiences, which were all sex-agnostic. Given the objective of this study, discussions are limited to sex-specific (sub-)themes and sex-agnostic themes that were reported differently between sexes.

### 4.1 Sex-Specific Prosthetic Experiences

Themes of Prosthetic Feel (1.1) and Prosthetic Function (1.2) emerged, relaying experiences of how the prosthesis feels on the body and how the prosthesis functions, respectively. When discussing Prosthetic Feel, females reported challenges with prosthetic fit, relaying how residual limb fluid volume and body fluctuations negatively impacted prosthetic fit (1.1.1). fLLPUs are often reported in literature, including in Phase-1, as having lower satisfaction and success with prosthetic fit,^12,28^ which can be attributable to many different factors including residual limb shape and volume fluctuations.^6^ Further, fLLPUs are more likely to experience greater fluid volume fluctuations than males.^44^ Therefore, this Prosthetic Feel theme offers a potential explanation for this female-specific experience. Further research may warrant consideration into the implications of sex-specific body fluctuations with prosthetic fit and satisfaction as well as the implications for sex-specific prosthetic design. Interestingly, a male-specific sub-theme within Prosthetic Feel centered around prosthetic weight, criticizing how heavy their device felt (1.1.2). This male-specific sub-theme is of interest as prosthetic weight is often a suggested reasoning in literature for why females have lower prosthetic satisfaction.^20,45,46^ While some female participants (n=3) reported dissatisfaction with prosthetic weight, it was not as prioritized a topic as it was for males. This could imply that males found their prosthetic weight more problematic than females. However, given the additional challenges reported by females regarding Prosthetic Feel, this could also imply there were points of greater consequence that females wished to discuss (e.g., shape and volume fluctuations) apart from prosthetic weight. Females are generally smaller and less strong than males,^47^ which likely limits successful prosthetic fit due to the componentry’s narrow selection for size and weight.^47^ However, it is worth considering and placing emphasis on further sex differences besides size and strength that may also be of consequence.

When discussing Prosthetic Function, females emphasized their priorities of ankle mobility (1.2.1), a theme unique to this study. Non-disabled (ND) females are known to have greater ankle ROM, exhibiting greater ankle plantarflexion in swing when compared to males.^48,49^ While prosthetic ankles are limited in their available ROM, it is possible fLLPUs are additionally limited in their mobility as their theoretical non-pathological gait would offer additional sagittal plane ROM, not accommodated for with generic prostheses. Further research should compare lower limb amputation and non-pathological gait biomechanics with a sex-specific lens to better understand gait compensations of fLLPUs and whether female-specific differences necessitate additional prosthetic ankle ROM or other sex-specific prosthetic design considerations.

### 4.2 Sex-Specific Physical Experiences

Sex-specific themes arose relating to Mobility (2.2) and Experiences with Pain (2.3). Physical mobility themes largely reinforce the literature narrative that fLLPUs experience greater challenges with ambulation and mobility,^8–12,50^ as well as novel findings from Phase-1.^28^ Female participants reported challenges ambulating on non-flat terrain, including walking on uneven ground and slopes (2.2.1). This female-specific sub-theme reinforces findings from Phase-1, where females reported a significantly poorer ability ambulating up or down a steep hill.^28^ Further, both females and males discussed having positive experiences with mobility (2.2.2), however, the experiences discussed were different. While females reported positive experiences with basic activities of daily living (e.g., walking short distances, getting up from a chair), males reported on significantly more advanced activities (e.g., walking significant distances, athletic activities). These sex-specific experiences further reinforce literature findings that females are more limited in their mobility.^8–12,28,50^ As well, these points emphasize the novel mobility themes derived in this work for further sex-specific consideration and study, which may not have emerged without adequate consultation with people of lived experience. Further research is needed to better understand these diverging experiences with mobility and what factors may be contributing. For instance, while tailored rehabilitation programs consider individual patient needs, they are not necessarily sex-specific.^7^ Future research should consider emphasizing sex-specific rehabilitation strategies to adequately target female mobility needs. Alternatively, it is possible that there are intrinsic sex differences in LLPUs’ gait biomechanics that create mobility disadvantages for fLLPUs. As mentioned, sex differences exist in ND gait biomechanics,^48,49,51^ and such differences are often thought to influence injury mechanisms in ND females.^51^ Unfortunately, there is no comparable research investigating sex differences in LLPUs’ gait biomechanics. Thus, research is warranted to elucidate whether such ND sex differences hold in LLPUs and how sex differences in LLPUs’ gait biomechanics across different terrains might play a role in sex-specific experiences with mobility as well as their implications for sex-specific rehabilitation and prosthetic design.

Lastly, both females and males reported Experiences of Pain (2.3), however, the severity of their reports were different. While females reported pain as manageable (2.3.2), male accounts were much more severe and intense (2.3.1). Reports and experiences of pain are largely influenced by both sex and gender,^52,53^ with biological influences relating to immune and hormonal factors as well as sociocultural influences relating to perceptions of gender identity.^53,54^ While analyses in this study have been limited to sex differences, interactions of sex and gender within the context of pain are difficult to distinguish, hence discussion drawing from both. Sex differences in hormonal factors and pain-related immune responses have been documented, suggesting females and males experience and process pain differently.^54^ From a gender-specific lens, women are more likely and willing to report pain, ^53–55^ while men are more willing to risk severe healthcare outcomes than seek medical attention, often to their detriment in preventative care.^53,56^ Women tend to seek a greater range of coping strategies than men, reported as feeling more responsible for managing their own pain and health.^53^ Furthermore, women are often thought to have greater body awareness and are thus more used to experiencing and managing pain.^53^ Overall, it is possible female participants in this study experienced pain that was more manageable, but it is also possible they sought out further medical attention than their male counterparts. And these possible explanations could stem from sex and/or gender-related differences. Regardless, findings from this study further contribute to the growing body of knowledge surrounding sex and gender differences in experiences of pain in the LLPU community, emphasizing the need for a sex and gender-specific lens to uncover these complex experiences.

### 4.3 Sex-Specific Psychosocial Experiences

Finally, sex-specific themes arose in psychosocial experiences, including through interactions with clinicians and life-style trade-offs. When discussing Frustrations with Medical Professionals (3.1), females reported not being taken seriously or listened to (3.1.1). This narrative mirrors literature findings, with females expressing that their health conditions are not taken seriously and that they feel mistrusted or psychologized by medical professionals.^20,53^ Such experiences can cause anxiety, which when articulated in tandem with pain or illness, results in a greater degree of psychologization.^53^ Furthermore, despite females reporting pain more often and seeking out more medical attention than males, males consistently receive more comprehensive medical treatments for the same diseases.^56^ For instance, males are thought to receive more effective pain relief treatment in the form of pain medication prescriptions while females are prescribed more antidepressants and received more mental health referrals in response to pain management decisions.^53^ While fLLPUs experience higher rates of mental health challenges and social burden,^9,28,57^ future research should investigate how negative experiences with clinicians might influence unique psychosocial challenges and contribute to feelings of distress, social burden, and/or mental health challenges.

Females also expressed themes relating to Life-Style Changes and Choices (3.2), particularly reporting new-found concerns around child-rearing (3.2.2). These triangulate and expand upon novel findings from Phase-1,^28^ where females reported their interest in having children being significantly affected because of their amputation. The qualitative findings from this study provide further depth from Phase-1,^28^ relating these concerns to maternity, including physical and psychosocial aspects of carrying a child to term, giving birth, and child-rearing. There is a lack of research addressing pregnancy in females with disabilities and existing research tends to neglect the unique perspectives of females with amputations within their population.^58^ Pregnancy requires significant physical and psychosocial adaptations over gestation that continue after birth, and the added effect of a limb difference presents further challenge.^58^ Moreover, this significant gap in literature on females with limb loss and pregnancy mirrors the lack of knowledge and experience medical professionals have in this space,^58^ all of which may contribute to feelings of hesitancy or uncertainty around aspects of maternity for fLLPUs. These findings of new concerns and uncertainty around having children are unique to this work, emphasizing the sex-specific gap in literature particularly as it relates to pregnancy in females with disabilities. Further, interview questions in this study relating to having children (formal or probing) were added through consultation with Patient Partners in efforts to better target their priorities with the interview guide. Thus, such themes may not have emerged without meaningful engagement with fLLPUs, demonstrating the immense benefit that can ensue by doing so. Further research is needed to investigate and understand these new-found concerns, what aspects of maternity are they stemming from, and how healthcare professionals can better support this population’s needs through pregnancy.

### 4.4 Limitations

Study limitations include cohort sampling and the small sample size. While purposive sampling methods may have been more appropriate to gain a wider range of perspectives, the study had to rely on convenience sampling methods due to the small population of LLPUs where the study was based, meaning the participant group consisted of those who responded to our recruitment methods. In efforts to diversify our cohort through a convenience strategy, various recruitment methods were used as outlined in Phase-1.^28^ Another sampling-related limitation is that all participants with TFAs in this study were male, limiting generalizability as findings may not necessarily extend across broader cohorts of fLLPUs. However, responses for participants with TTAs versus TFAs did not markedly differ in this study. As well, there were no statistically significant differences between sexes for any demographic parameter, including amputation level, making these good comparative groups. Overall, despite sampling limitations, code saturation was achieved across participants (across sexes, amputation levels, etc…), and many findings were triangulated with Phase-1,^28^ and followed similar trends from literature.

As discussed, since the cohort comprised only of cisgender people, there were not enough gender-diverse perspectives to disambiguate sex versus gender differences. Therefore, (sub-)themes were considered as either sex-specific or sex-agnostic. However, particularly in the context of pain and healthcare experiences, the authors acknowledge the complex interactions between sex and gender. Therefore, discussion of the existing literature drew from both concepts to elucidate potential explanations.

Overall, research addressing sex differences in LLPUs is lacking. This exploratory study was meant to expand initial insights and begin uncovering and understanding sex differences in LLPUs. Future research with a more gender-diverse cohort may seek to expand findings and the presented explanations to reclassify within a sex and/or gender context. As well, findings should be expanded with a wider range of perspectives through purposive sampling methods, considering females with TTAs and TFAs to explore their unique experiences.

## 5. Conclusion

This study utilized POR to develop a semi-structured interview guide that better targets key priorities of fLLPUs. Interview findings present an array of sex-specific experiences in prosthetic, physical, and psychosocial aspects, expanding understanding of sex differences in the perspectives, priorities, and lived experience of LLPUs. Furthermore, findings present new avenues and insight for future research to address the unique needs of fLLPUs through sex-specific considerations in rehabilitation and prosthetic design.

## Declaration of interest

The authors report no conflicts of interest.

## Supporting information

SDC-1 Codebook

SDC-2 Table 1S

## Data Availability

All data produced in the present work are contained in the manuscript.

## Acknowledgments

This study was made possible by members of the Patient-Oriented Research committee, including Julia Chisholm, L. Hall, Jessica Nathan, Diane Reid, and Gail Timmer. These members provided invaluable insight into experiences and perspectives of female lower limb prosthesis users and collaboratively worked with the authors to outline objectives for this study and co-build the interview guide.

Graduate funding for TMRC was provided by the British Columbia Graduate Scholarship (BCGS), the BC SUPPORT Unit Patient-Oriented Research Fellowship, NSERC Canada Graduate Scholarships – Doctoral (CGS-D), and CIHR-IMHA Inclusive Research Excellence Prize for Patient Engagement. These sources of funding had no input on the study design nor on the collection, analysis and interpretation of data, or manuscript preparation. They also had no input on the preparation of the manuscript nor on the decision to submit the manuscript for publication.

## Notes

### Competing Interest Statement

The authors have declared no competing interest.

### Author Declarations

The Research Ethics Board of the University of Victoria and the Vancouver Island Health Authority gave ethical approval for this work (H21-00778).

