## Supplementary material for "A Qualitative Study Investigating Sex Differences in the Needs and Priorities of Lower Limb Prosthesis Users": SDC-1 Codebook

### Supplemental Digital Content 1

##### Finalized Codebook

| **#** | **Code** | **Subcode level 1** | **Subcode level 2** | **Description** | **Line #s** | **Example** |
| --- | --- | --- | --- | --- | --- | --- |
| **1** | **Prosthesis** | | | | | |
| 1.1 |  | Prosthesis weight |  | Text coded to topics of discussion on weight of someone's prosthesis or prosthesis components. | INT003: 15-17 | INT003: "I am very active, and I think there's two things that limit me with this prosthetic, and one of them is the weight of the actual prosthetic, and the other one is the difficulty in maintaining a real physical level of activity without sweating, and having to reset and take it off numerous times when I’m active." |
| 1.1.1 |  |  | Lightweight | Text coded to topics of discussion of their prosthesis (or components) being too heavy or desiring their prosthesis to be lighter weight. | INT003: 374-380 | INT003: "My ideal prosthetic would be very light. It would probably be made of… bamboo. I’m not sure, there's a guy in India that actually makes them because they needed prosthetics that were cheap, and you know, and, and that they could, that they could share with people that didn't have access to them. He was making prosthetics out of bamboo. And so not only were they really light, but they were also from this renewal, like this crazy, renewable source, right? That and I just thought, man, that's awesome, that's so great. So I’m not saying I would actually like one but I’d like the idea of having a very very light one." |
| **#** | **Code** | **Subcode level 1** | **Subcode level 2** | **Description** | **Line #s** | **Example** |
| 1.2 |  | Prosthesis adjustability |  | Text coded to topics of discussion around the interest or desire to have more adjustability, range of motion, and/or flexibility with a prosthesis or being frustrated with these aspects of their own prosthesis, prosthesis' ability to accommodate different shoes/clothes, prosthesis' ability to accommodate body's variability day to day or season to season. | INT004: 355-359 | INT004: "Ideally, I'd like to have an ankle that moved. Um… Had a little flex side to side. My foot does have a little bit flex but, I think rough ground it’d benefit from more, more flex. Um, a heel on the foot, also that's, would be adjustable. So right now, I gotta wear shoes that are almost the same from heel to toe. I, we call it the heel-drop. Now, if I wear something else, it's got more of a heel in it, like a work boot or something, it tips me forward. " |
| 1.2.1 |  |  | Ankle adjustability | Text coded to topics of discussion around the interest or desire to have more adjustability, range of motion, and/or flexibility with the foot/ankle components of the prosthesis, or being frustrated with these aspects of their own prosthesis. Desires could include having adjustable heel height or be able to perform more motions. Prosthesis' ability to accommodate different shoes/clothes. | INT003: 382 | INT003: "I’d like the option of having that adjustable heel. " |
| 1.2.2 |  |  | Socket and suspension adjustability | Text coded to topics of discussion around the interest or desire to have more adjustability, range of motion, and/or flexibility with the socket, or being frustrated with these aspects of their own prosthesis. Desires could be around wanting to adjust it on the go, having easy/quick adjustability. Prosthesis' ability to accommodate body's variability day to day or season to season. | INT004: 365-369 | INT004: "Um… and then a socket that's more flexible. Yeah. Move around, some adjustability. So, uh, one day, if I eat a bunch of potato chips or whatever, you know, the leg’s gonna be swollen up. And okay, well, I gotta put less socks on… the liner for fit. And then I go to the gym and sweat a bit, and then all of sudden, it's like too small and it's twisting around on me all the time. So adjustability. Just, I think that would be a game changer. Big time." |
| **#** | **Code** | **Subcode level 1** | **Subcode level 2** | **Description** | **Line #s** | **Example** |
| 1.3 |  | Prosthesis fit / comfort |  | Text coded to topics of discussion around perceived fit of the prosthesis, satisfaction with this, how the prosthesis is fitting (good or bad), the affect of prosthesis fit, the technology used to fit it, wearability and comfort of the prosthetic, any mention of comfort/discomfort wearing the prosthesis. | INT003: 79-80 | INT003: "Prosthetic fit is a huge part of that mobility and comfort is what enables me to do what I do everyday." |
| 1.3.1 |  |  | Poor fit | Text coded to topics of discussion around a prosthesis not fitting well or having low satisfaction with the fit or wishing for improvements on the fit, causing pressure sores, or causing twist/skin shear, discussion of prosthesis being uncomfortable or causing discomfort | INT005: 28-30 | INT005: "Um… I think it starts with the fit. So the fit is not proper. And I think with that, then comes like certain alignment issues. Right? So it's not fitting properly. So it's causing sores, discomfort, and then yeah, causing me to walk out of alignment, etc." |
| 1.3.2 |  |  | Prosthesis technology | Text coded to topics of discussion around the technology used to fit prosthesis, used to create custom sockets, satisfaction with technologies and the fitting process, time required to make the prosthesis or get repairs. Also, technology used to create the prosthesis, new prosthesis technology. Overall, technology for prosthesis fit and function. | INT003: 60-62 | INT003: "It would be nice to see more modern approaches to fitting a prosthetic such as 3D scanning. I’m curious about that, and I’m curious about what would be available if funding was more fair for individuals." |
| 1.3.3 |  |  | Impacts of prosthesis fit | Text coded to topics of discussion around the effect prosthesis has on different aspects of life, e.g., comfort, mobility, mood etc…, causing damage or pain to the residual limb. Can cause skin shearing if too tight or twisting if too loose. Can affect joint or back pain/problems | INT003: 79-80  INT005: 28-30 | INT003: "Prosthetic fit is a huge part of that mobility and comfort is what enables me to do what I do everyday."  INT005: "Um… I think it starts with the fit. So the fit is not proper. And I think with that, then comes like certain alignment issues. Right? So it's not fitting properly. So it's causing sores, discomfort, and then yeah, causing me to walk out of alignment, etc."" |
| **#** | **Code** | **Subcode level 1** | **Subcode level 2** | **Description** | **Line #s** | **Example** |
| 1.4 |  | Prosthetic trade-offs |  | Text coded to topics of discussion around trade-offs experienced with prosthetics, when the prosthetic tries to target one attribute while negatively impacting another attribute or needing multiple legs to target different activities | INT008: 98-107 | INT008: "Um like the for example, the reason why I actually use my skiing for riding is because of the angle of the heel joint essentially, is adjusted differently than my walking foot, so because when you're mounted I mean you're in a different position and with my regular foot and it's just it, the angle sits at that makes it difficult to post while I’m trotting to go stand up into like standing up in the stirrups you know things like that. But when I’m on the ground that same foot isn't good for walking, because of that angle you know it-it's-and also the fact that it has a it designed to for clicking into a ski. So it has the jut out from the heel. So that you know it makes it hard to walk, because, you know, you have an extra 3 quarters of an inch further back from where you, where your heel would normally be. So it changes your gait, it's like it's just like walking in a ski boot." |
| 1.4.1 |  |  | Mobility vs stability | Text coded to topics of discussion around trade-offs experienced with prosthetics regarding mobility versus stability. A more stable prosthetic does not allow for as much range of motion and flexibility and thus does not allow as much movement or mobility and vice versa. Discussions around this topic. | INT002: 54-58 | INT002: "When it comes to things like cycling, it's really difficult because I don't have enough of a cutout in the back of my prosthetic to allow the proper movement, and I can’t because then I lose stability when trying to walk. This means I would require a second socket altogether, one made only for cycling. So, that's really frustrating." |
| 1.4.2 |  |  | Needs vs Afford | Text coded to topics of discussion around trade-offs between a prosthetic that meet your needs and one that you can afford. How getting the right prosthetic for you, one that meets your needs and is all the things you want is too expensive. There are trade-offs with the ideal prosthetic/what it can do and accomplish and what someone can actually afford or the idea of trade-offs existing between getting all the prosthetics you want/need for your lifestyle and managing what you can afford | INT002: 72-76 | INT002: "Feet with more spring and a natural ankle motion, greatly improve mobility and there are prosthetic feet like this, but they have their own issues. No foot is perfect. These feet tend to be electrical or mechanical which means they require a lot of maintenance over the years. It's something that you use for up to 14 hours a day, so there can be a lot of wear and tear. The other issue is that they cost a lot. For instance, the one that I was going to use was around $30,000. So I decided against that." |
| **#** | **Code** | **Subcode level 1** | **Subcode level 2** | **Description** | **Line #s** | **Example** |
| 1.4.3 |  |  | Aesthetics vs function | Text coded to topics of discussion around trade-offs experienced with prosthetics regarding aesthetics versus function. A more "aesthetically pleasing" prosthesis typically is less functional or less practical and vice versa. Discussions around this topic. | INT003: 94-97 | INT003: "Aesthetics are important too. I just found that over so many years, I was willing to hide it and just settle for what was available. And there weren't many options at all. It was either a pylon look or a foam covering. Trying to match your own skin tone and muscle shape, it never looked quite right as best as they tried." |
| 1.4.4 |  |  | Jack of all trades prosthetic | Text coded to topics of discussion around trade-offs and limitations experienced with prosthetics and the activities they can accommodate. Topics around the idea of needing or using different prosthetics to perform different activities and there isn't a jack of all trades prosthetic (one that can do everything). | INT008: 109-111   INT002: 59-60 | INT008: "So basically, when I go riding I’m, I’m wearing my regular leg carrying in my ski leg in the arena then when I go up to the mounting block, I need to sit down, swap out the leg, climb the mounting block, and get on the horse, you know. "  INT002: "The fact that I require so many different types of prosthetic feet to do the things that I want to do is pretty frustrating." |
| 1.5 |  | Attitude toward prosthesis |  | Text coded to topics of discussion around someone's attitude or feelings towards their prosthesis, whether these are positive or negative. This could relate to feelings of frustration about a prosthesis, or feelings of being satisfied. This could also be coded for the mindset someone has about their prosthesis, like that their prosthesis is a personal device, or that their prosthetic makes them feel whole or complete. | INT010: 281-283 | INT010: "And yeah, [my prosthesis is] a very personal item, it’s an extension of myself so when I don’t wear my leg, I don't feel whole like if that makes sense like, I know I removed part of my body, and this is me naturally, but I feel more whole with this artificial limb." |
| **#** | **Code** | **Subcode level 1** | **Subcode level 2** | **Description** | **Line #s** | **Example** |
| **2** | **Physical** | |  |  |  |  |
| 2.1 |  | Energy Required |  | Text coded to topics of discussion around perceived exertion to complete daily tasks or perceived energy required to perform daily activities, how tired an activity made someone. | INT003: 30-32 | INT003: "But for myself, as a woman I'd be super happy if I had a really light prosthetic. Because it takes a lot of physical energy to use a prosthetic and when you add heavier components, you expend more energy to utilize them. " |
| 2.1.1 |  |  | Physical Exertion | Text coded to topics of discussion around the perceived physical exertion of performing daily tasks, saving energy to perform basic tasks (e.g., accessible parking when going to a store), this could be related to prosthesis weight or muscle tiredness or skin discomfort or joint discomfort etc... | INT003: 30-32 | INT003: "But for myself, as a woman I'd be super happy if I had a really light prosthetic. Because it takes a lot of physical energy to use a prosthetic and when you add heavier components, you expend more energy to utilize them. " |
| 2.1.2 |  |  | Mental Exertion | Text coded to topics of discussion around the perceived mental exertion of performing daily tasks, always having to think about the next step or move, having to focus on your movements and being aware of your surroundings. | INT003: 435-440 | INT003: "It takes a lot of energy, just no matter what you're doing, as an amputee. It just takes more energy and a lot more thought process, even when walking. When I’m walking in the forest, I’m looking around all the time. I’m perceiving the little bumps in the ground, the proprioception is not there on the one side so you're always, you're always mentally scanning things, perceiving ways to navigate. A lot of people walking will be looking at buildings, looking at the sky, looking at, you know, nature, the forest, the trees. But I’m always looking down, looking around." |
| **#** | **Code** | **Subcode level 1** | **Subcode level 2** | **Description** | **Line #s** | **Example** |
| 2.2 |  | Residual Limb Attributes |  | Text coded to topics of discussion around someone's residual limb, the state and condition of that residual limb (e.g., is it sore with a lot bumps/calluses?, is there pain caused by neuromas), the length of the residual limb, the shape/size of the limb, and the effect of these factors on day to day life/activities, effect on prosthesis fit or satisfaction, etc... | INT005: 134-137   INT005: 405-408 | INT005: "Well, there's definitely another issue at play and it is my limb. So throughout this whole thing, I’ve been told that I am a unique case in the sense that my amputation of my leg was not textbook. So there's a ton of scar tissue, there's different tissue down there, a lot of skin graft, a lot of scar tissue, etc. So I know that complicates the situation."  INT005: "I think it's a combination of both, like there are some spots there where yeah, it's like blisters, open sores. But then I also think it's just pressure from the socket, like, like, just yeah, just pushing on spots. So I think it's a combo of the fit and the sores that are there. " |
| 2.2.1 |  |  | Residual limb length | Text coded to topics of discussion around the effect of limb length on various aspects of their daily life, including mobility, prosthesis fit, general satisfaction. | INT002: 8-10 | INT002: "It's difficult for me to truly know because I have a shorter limb and neither my surgeon nor my prosthetist explained how that would affect me but I do believe it hinders my ability to have a well-fitting prosthetic." |
| 2.2.2 |  |  | Wound care | Text coded to topics of discussion around wounds on the residual limb needing care and attention from medical professionals | INT010: 150-155 | INT010: "They debride it every now and then, and it heals up rapidly to a certain point, and then it stagnates. Then they debride it again. And they know that like- I I just took 6 months off work. Yeah. So now, but back in, back doing you know back into the workforce, the wound care team knows that I have to work, and that I am gonna work. So it's just a matter of maintaining those wounds and the pain until you know winter comes and then off work again." |
| 2.2.3 |  |  | Residual Limb Health | Text coded to topics of discussion around health of residual limb, if sores are present, if skin is irritated, if there are places were the bone is rubbing into the skin, and how this affects day to day life/activities and comfort with prosthesis, fit of prosthesis, scar tissue, etc... Are they dealing with neuromas or similar pain related to limb health, i.e., not just skin challenges but health below the skin's surface too. | INT005: 134-137 | INT005: "Well, there's definitely another issue at play and it is my limb. So throughout this whole thing, I’ve been told that I am a unique case in the sense that my amputation of my leg was not textbook. So there's a ton of scar tissue, there's different tissue down there, a lot of skin graft, a lot of scar tissue, etc. So I know that complicates the situation." |
| **#** | **Code** | **Subcode level 1** | **Subcode level 2** | **Description** | **Line #s** | **Example** |
| 2.3 |  | Body fluctuations |  | Text coded to topics of discussion around weight fluctuations in the residual limb or of the overall body, frustrations around this, effect of this on daily life activities like prosthesis comfort/mobility/overall satisfaction/fit. Also discussion around changes in shape of the limb or body (i.e., not just size but shape too) and how this affects day to day life. | INT002: 199-200 | INT002: "It's mostly just you know how easy is it going to be for me to carry a baby. How easy is it because of weight fluctuations, especially with swelling and stuff." |
| 2.3.1 |  |  | Swelling of amputated limb | Text coded to topics of discussion around swelling of the residual limb, frustrations around this and impacts of this on daily life as well as suggestions to manage or improve it, what triggers it as well | INT004: 365-369 | INT004: "Um… and then a socket that's more flexible. Yeah. Move around, some adjustability. So, uh, one day, if I eat a bunch of potato chips or whatever, you know, the leg’s gonna be swollen up. And okay, well, I gotta put less socks on… the liner for fit. And then I go to the gym and sweat a bit, and then all of sudden, it's like too small and it's twisting around on me all the time. So adjustability. Just, I think that would be a game changer. Big time. " |
| 2.3.2 |  |  | Shrinking of amputated limb | Text coded to topics of discussion around shrinking of the residual limb, frustrations around this and impacts of this on daily life as well as suggestions to manage or improve it, what triggers it as well | INT004: 365-369 | INT004: "Um… and then a socket that's more flexible. Yeah. Move around, some adjustability. So, uh, one day, if I eat a bunch of potato chips or whatever, you know, the leg’s gonna be swollen up. And okay, well, I gotta put less socks on… the liner for fit. And then I go to the gym and sweat a bit, and then all of sudden, it's like too small and it's twisting around on me all the time. So adjustability. Just, I think that would be a game changer. Big time. " |
| 2.4 |  | Sweat |  | Text coded to topics of discussion on someone being sweaty or their prosthesis being sweaty, their prosthesis making them too warm, their prosthesis making them sweaty, how sweat or feeling too warm can affect different things (e.g., loosening liner/fit/comfort/satisfaction/noises). | INT003: 380-382 | INT003: "I like the idea of not having so many layers. I think when, by the time I don mine, I’ve got seven layers next to my skin and, and that contributes a lot the sweat too. So yeah, I thought, I’d love that." |
| **#** | **Code** | **Subcode level 1** | **Subcode level 2** | **Description** | **Line #s** | **Example** |
| 2.5 |  | Pain |  | Text coded to topics of discussion around pain of any type relating to their amputation endured during daily life activities, including frustrations with the pain, what triggers it, suggestions of how it can be managed or improved | INT001: 4-5 | INT001: "Umm with my prosthesis, yes. [My certified prosthetist] is extremely good, but for me it's not the prosthesis, it's my leg itself. It, my leg has been, for the last year, in so much pain. " |
| 2.6 |  | Mobility |  | Text coded to topics of discussion around all things mobility. Discussions of desired mobility levels (i.e., walking vs running vs cycling), lacking mobility, what mobility means to them, how could mobility be improved, etc… | INT003: 78-79 | INT003: "Sure, priority for me as I said before is mobility, it's walking, it's being able to get from A to B without struggle. It's being able to take part in my life the way I want to." |
| **#** | **Code** | **Subcode level 1** | **Subcode level 2** | **Description** | **Line #s** | **Example** |
| **3** | **Psychosocial** | |  |  |  |  |
| 3.1 |  | Mental Health |  |  |  |  |
| 3.1.1 |  |  | Peer Support | Text coded to topics of discussion around the importance of peer connection and supports, having a community or team, having a family, and the importance of the support from others, being social with others. or having a lack of peer support. To note, this support is from close peers (i.e., family, friends) but not necessarily from support groups or support from professionals | INT009: 401-403 | INT009: "I had a really an exceptional support mechanism around like my network of family and friends are through the roof for me. Yeah, that's what got me through all that more so than any, you know, professional support." |
| 3.1.2 |  |  | Social Acceptability and  Self-image | Text coded to topics of discussion around desiring things about oneself to meet social norms or be socially acceptable, like how one walks or how one dresses, topics around how you are perceived socially (by others), desire to not draw attention to oneself or maybe the opposite. This all pertains to the idea of "social-perception" and how others see you. Or discussion around someone's self-image (i.e., how someone views or perceives themselves) and how it has (or has not) changed after amputation, how it changes (if at all) when wearing a prosthetic. This all pertains to the idea of "self-perception" and how you see you. | INT008: 250-254  INT008: 261-264 | INT008: "...sometimes like when my prosthetic isn't visible and I use a handicap parking stall, there's times that I feel like people look at me and think I’m a fraud, you know, because I mean, when I’m wearing pants, my missing leg is a hidden disability. So that happens once in a while, just you know, like are people looking at me and judging me, or you know, sometimes it happens to yourself."  INT008: "...sometimes you get into your own head like “Should I be doing this?” you know, like, “Should I be using this stall, or should I be leaving it available for someone else?” know stuff like that. And so I mean it does play a role in and it does lead to that whole “Do you view yourself as being disabled or handicapped or not?" |
| **#** | **Code** | **Subcode level 1** | **Subcode level 2** | **Description** | **Line #s** | **Example** |
| 3.1.3 |  |  | Grieving | Text coded to topics of discussion around needing to grieve the loss of a limb, grieving the life lost, grieving loss of a career, grieving the loss of acceptance in social settings, grieving the life changes | INT009: 385-387 | INT009: "I went through all the stages of grief that everyone does with limb loss, which is, you know, the same stages that you go through when you're grieving anything else." |
| 3.1.4 |  |  | Fears | Text coded to topics of discussion around being afraid to complete day to day activities (e.g., using the oven/baking) or fear of falling or fear of doing activities alone, fear of living alone and how that would work with their needs. Also discussions of general concerns, worries, or stresses of performing activitIes, not just the feeling of fear. | INT013: 117-118 | INT013: "But there is that- I think you know I still have a fear of falling. Yeah, I think that's partly what it's about fear of falling." |
| 3.1.5 |  |  | Attitude/ Mindset | Text coded to topics of discussion around how your mindset or attitude towards things can greatly affect your satisfaction and quality of life. | INT008: 270-271 | INT008: "You know, so it's all about the mindset of having things that limit you versus having things that empower you, you know." |
| 3.2 |  | Independence |  |  |  |  |
| 3.2.1 |  |  | Ability to Drive | Text coded to topics of discussion around the ability to drive and the impact of this ability on independence, confidence required when driving/to-relearn. | INT001: 144-146 | INT002: "I find now that I can't drive, I can’t enjoy the audience I used to enjoy. You can't meet the group for dinner somewhere. You can't pop into a grocery store. And with a prosthetic on, you can't go in and try on clothes at a store." |
| **#** | **Code** | **Subcode level 1** | **Subcode level 2** | **Description** | **Line #s** | **Example** |
| 3.3 |  | Medical professionals and services |  |  |  |  |
| 3.3.1 |  |  | Frustrations and limitations with medical professionals | Text coded to topics of discussion around limitations and frustrations experienced with family doctors/general physicians, GPs not having worked with amputees/having very little training with amputees. This can also be frustrations with medical professionals not having enough time for appointments. Also topics of requiring to self-advocate for oneself about their health, around possible frustrations of being taken seriously or listened to by medical professionals, self-educate and can't rely on medical professionals to some degree, self-educate about prosthesis types/what would be suitable for oneself, and around the communication or lack-there-of between medical professionals and their patients. | INT003: 204-209 | INT003: "Having said that, I've had to teach many health care professionals that I’ve come across over the years about my needs because they don't have that background, or they haven’t experienced it. And I’m still running into that, you know, 30 years later. In the last 5 years I’ve been dealing with the issues with sweat with the prosthetic. More so teaching a dermatologist about possible options that are out there to resolving it. So I’m always having to advocate on some level for… and educate on some level. " |
| 3.3.2 |  |  | Programs and services | Text coded to topics of discussion around training programs and/or structured support services, needing more of these services or having had a positive or negative experience with them. Supports/training before, during, and after the amputation (longitudinal). This could be through structured support groups or through medical professionals offering gait training, etc... | INT009: 372-376 | INT009: "[Staff at the rehab facility] on the, you know, they're on the timer to say, “this person is supposed to be finished in 4 weeks, and when he's done, he's done”. So uh, I wish there was a secondary follow up program involved that where somebody phones me 2 years later, and said “Hey? how's it going? Do you want to come back in for a bit of a revisit?” That would just be wonderful if someone did that, but that doesn't happen." |
| **#** | **Code** | **Subcode level 1** | **Subcode level 2** | **Description** | **Line #s** | **Example** |
| 3.4 |  | Lifestyle Trade-offs |  | Text coded to topics of discussion around any trade-offs experienced in life post-amputation. Things like career trade-offs, financial trade-offs, personal trade-offs (e.g., having children), mobility/activity trade-offs, clothing trade-offs, other daily life trade-offs, etc... | INT002: 146-153 | INT002: "I think it has. I was never one that felt like I needed children in my life to begin with, but it has made it a lot harder to want children, imagining myself as an amputee-mother. My husband's had to help me out so much as it is, and the thought of him having to take on a baby as well is a lot. If anything goes wrong with me, especially since complications with my amputation seem to continuously arise, it scares me that he would have to deal with so much- work and finances included. Getting up in the middle of the night, trying to balance the cost of being an amputee and having a baby, imagining myself keeping up with, or having enough energy to care for a child, dealing with the extra weight during and after pregnancy…that kind of stuff is difficult." |
| **#** | **Code** | **Subcode level 1** | **Subcode level 2** | **Description** | **Line #s** | **Example** |
| **4** | **Finances** | |  |  |  |  |
| 4.1 |  | Amputee expenses |  | Text coded to topics of discussion on expenses incurred as an amputee, including cost of prostheses, cost of living, cost of other medical expenses as an amputee… | INT002: 151-153 | INT002: "Getting up in the middle of the night, trying to balance the cost of being an amputee and having a baby, imagining myself keeping up with, or having enough energy to care for a child, dealing with the extra weight during and after pregnancy…that kind of stuff is difficult. " |
| 4.1.1 |  |  | Prosthesis affordability | Text coded to topics of discussion around the affordability/cost of prosthetics (and components/liners/compression socks), cost of prosthesis repairs, etc… cost of having multiple prosthetics or having high functioning/quality ones, or sport/activity specific prosthetics, prosthetic warranty (i.e., through the prosthetic manufacturer and what they cover if things break) | INT002: 72-76 | INT002: "Feet with more spring and a natural ankle motion, greatly improve mobility and there are prosthetic feet like this, but they have their own issues. No foot is perfect. These feet tend to be electrical or mechanical which means they require a lot of maintenance over the years. It's something that you use for up to 14 hours a day, so there can be a lot of wear and tear. The other issue is that they cost a lot. For instance, the one that I was going to use was around $30,000. So I decided against that." |
| 4.1.2 |  |  | Insurance and coverage | Text coded to topics of discussion around insurance coverage on prosthetics or on other life expenses related to being an amputee (e.g., wheelchairs, crutches, bathtub seats - anything like this), insurance companies (Pharmacare), what insurance companies deem as a "necessity" or deem as "basic", etc... | INT003: 64-74 | INT003: "Pharmacare is my first go-to. I’m lucky that I have extended health insurance as well but often that doesn't even cover needs. Extended health coverage lacks understanding of the realities of amputee life and how certain components are necessary to fit a prosthetic leg. For example, they will not cover the liners that I need to actually don/wear my leg. Pharmacare will cover two a year but when you alternate two every day, all day, all year, it's similar to asking a person to wear the same pair of socks for a year. " |
