## Supplementary material for "A Qualitative Study Investigating Sex Differences in the Needs and Priorities of Lower Limb Prosthesis Users": SDC-2 Table 1S

**Table 1S.** Summary of demographics for each recruited participant.

| **ID** | **Age Range* (years)** | **BMI (kg/m^2^)** | **Sex Assigned at Birth** | **Gender Identity** | **Amputation Level** | **Time Since Amputation (years)** | **Time with Current Prosthesis (years)** | **Amputation Pathology** | | **Prosthesis Type**   - **Suspension** - **Knee** - **Ankle** |
| --- | --- | --- | --- | --- | --- | --- | --- | --- | --- | --- |
| 1 | 75-79 | 28.4 | Female | Woman | TTA | 1-2 | <1 | Vascular | | Undisclosed |
| 2 | 30-34 | 22.2 | Female | Woman | TTA | 1-2 | <1 | Cancer | | - Undisclosed - N/a - Ossur Pro-Flex LP Align Foot |
| 3 | 45-49 | 19.9 | Female | Woman | TTA | >10 | <1 | Trauma | | - Elevated Vacuum - N/a - Ossur Pro-Flex Foot |
| 4 | 50-54 | 29.7 | Male | Man | TFA | 1-2 | <1 | Trauma | | - Lanyard System - Ottobock Genium X3 Knee - Ottobock Taleo Foot |
| 5 | EXCLUDED - INELIGIBLE | | | | | | | | | |
| 6 | 60-64 | 32.3 | Male | Man | TTA | >10 | 1-2 | Trauma | | - Elevated Vacuum - N/a - Pro-Flex Pivot Foot |
| 7 | EXCLUDED - INELIGIBLE | | | | | | | | | |
| 8 | 65-69 | 23.2 | Male | Man | TFA | >10 | >5 | Trauma | | - Undisclosed - Ottobock Genium Knee - Ossur Pro-Flex LP Foot |
| 9 | 40-44 | 30.8 | Male | Man | TTA | 3-5 | 1-2 | Other | | - Pin Lock - N/a - Freedom Maverick Low Profile Foot |
| 10 | 30-34 | 20.8 | Female | Woman | TTA | 3-5 | <1 | Trauma | | - Pin Lock - N/a - Soleus Foot by College Park |
| 11 | 70-74 | 24.1 | Male | Man | TFA | >10 | 3-5 | Trauma | | - Undisclosed - C-leg Knee - Undisclosed |
| 12 | 75-79 | 26.6 | Female | Woman | TTA | 1-2 | <1 | Vascular | | - Pin Lock - N/a - Undisclosed |
| 13 | 60-64 | 20.8 | Male | Man | TTA | 6-10 | 3-5 | Trauma | | Undisclosed |
| 14 | 20-24 | 35.2 | Female | Woman | TTA | >10 | 3-5 | Other | | - Undisclosed - N/a - Pro-Flex Loop Integrated Foot |
| 15 | 55-59 | 30.5 | Male | Man | TFA | >10 | 3-5 | Vascular | | - Suction Socket - Blatchford Knee unit, DeltaTwist - Undisclosed |
| 16 | 35-39 | 30.3 | Female | Woman | TTA | >10 | 3-5 | Other | | Undisclosed |
| 17 | 70-74 | 23.6 | Female | Woman | TTA | 3-5 | 1-2 | Trauma | | - Suction Socket - N/a - Ossur Pro-Flex LP Align Foot |
| 18 | 85-89 | 21.9 | Female | Woman | TTA | >10 | 3-5 | Vascular | | Undisclosed |
| 19 | 75-79 | 32.5 | Male | Man | TFA | 6-10 | 3-5 | Vascular | | - Undisclosed - Ottobock Kenevo Microprocessor Knee - Ottobock Foot |
| 20 | 70-74 | 28.1 | Male | Man | TTA | 1-2 | <1 | Other | | - Pin Lock - N/a - Undisclosed |

Abbreviations: SD, standard deviation; BMI, body mass index; TTA, transtibial amputee; TFA, transfemoral amputee.

*Note: While precise ages were collected from participants in the study, only age ranges are presented here to preserve participant anonymity.

Note: While geographic location was collected, those information is not included here for anonymity purposes.
